# AI-Derived Splenic Response in Cardiac PET Predicts Mortality: A Multi-Site Study

**DOI:** 10.1101/2025.06.27.25330278

**Authors:** Naga L Dharmavaram, Giselle Ramirez, Aakash Shanbhag, Robert JH Miller, Paul B. Kavanagh, Jirong Yi, Mark Lemley, Valerie Builoff, Anna Marcinkiewicz, Damini Dey, Jon Hainer, Samuel Wopperer, Stacey Knight, Viet T. Le, Steve Mason, Erick Alexanderson, Isabel Carvajal-Juarez, René RS Packard, Thomas L. Rosamond, Mouaz H. Al-Mallah, Leandro N. Slipczuk, Mark I. Travin, Wanda Acampa, Andrew J. Einstein, Panithaya Chareonthaitawee, Daniel S. Berman, Marcelo F. Di Carli, Piotr J. Slomka

**Affiliations:** Departments of Medicine (Division of Artificial Intelligence in Medicine), Imaging, Cardiology, and Biomedical Sciences, Cedars-Sinai Medical Center, Los Angeles, California, USA; Department of Electrical and Computer Engineering, University of Southern California, Los Angeles, CA, USA; Department of Cardiac Sciences, University of Calgary, Calgary Alberta, Canada; Center of Radiological Diagnostics, National Medical Institute of the Ministry of the Interior and Administration, Warsaw, Poland; Division of Cardiovascular Medicine, Department of Medicine, Brigham and Women’s Hospital, Harvard Medical School, Boston, MA, USA; Department of Cardiovascular Medicine, Mayo Clinic, Rochester, MN, USA; Intermountain Medical Center Heart Institute, Intermountain Healthcare, Murray, UT, USA; Department of Internal Medicine, University of Utah, Salt Lake City, UT, USA; Department of Nuclear Cardiology, Ignacio Chavez National Institute of Cardiology, Mexico City, Mexico; Faculty of Medicine, National Autonomous University of Mexico, Mexico City, Mexico; Division of Cardiology, Department of Medicine, David Geffen School of Medicine, University of California, Los Angeles, CA, USA; West Los Angeles Veterans Affairs Medical Center, Los Angeles, CA, USA; Department of Cardiovascular Medicine, The University of Kansas Medical Center, Kansas City, KS, USA; Houston Methodist Academic Institute, Weill Cornell Medicine, Houston Methodist DeBakey Heart and Vascular Center, Houston, TX, USA; Cardiology Division, Montefiore Medical Center, Albert Einstein College of Medicine, Bronx, NY, USA; Department of Radiology, Montefiore Medical Center and the Albert Einstein College of Medicine, Bronx, NY, USA; Department of Advanced Biomedical Sciences, University of Naples Federico II, Naples, Italy; Division of Cardiology, Department of Medicine, and Department of Radiology, Columbia University Irving Medical Center and New York-Presbyterian Hospital, New York, NY, USA

**Keywords:** Artificial Intelligence, Splenic Switch-off, Nuclear Cardiology, Myocardial perfusion imaging

## Abstract

**Background:** Inadequate pharmacologic stress may limit the diagnostic and prognostic accuracy of myocardial perfusion imaging (MPI). The splenic ratio (SR), a measure of stress adequacy, has emerged as a potential imaging biomarker.

**Objectives:** To evaluate the prognostic value of artificial intelligence (AI)-derived SR in a large multicenter ^82^Rb-PET cohort undergoing regadenoson stress testing.

**Methods:** We retrospectively analyzed 10,913 patients from three sites in the REFINE PET registry with clinically indicated MPI and linked clinical outcomes. SR was calculated using fully automated algorithms as the ratio of splenic uptake at stress versus rest. Patients were stratified by SR into high (≥90th percentile) and low (<90th percentile) groups. The primary outcome was major adverse cardiovascular events (MACE). Survival analysis was conducted using Kaplan-Meier and Cox proportional hazards models adjusted for clinical and imaging covariates, including myocardial flow reserve (MFR ≥2 vs. <2).

**Results:** The cohort had a median age of 68 years, with 57% male patients. Common risk factors included hypertension (84%), dyslipidemia (76%), diabetes (33%), and prior coronary artery disease (31%). Median follow-up was 4.6 years. Patients with high SR (n=1,091) had an increased risk of MACE (HR 1.18, 95% CI 1.06–1.31, p=0.002). Among patients with preserved MFR (≥2; n=7,310), high SR remained independently associated with MACE (HR 1.44, 95% CI 1.24–1.67, p<0.0001).

**Conclusions:** Elevated AI-derived SR was independently associated with adverse cardiovascular outcomes, including among patients with preserved MFR. These findings support SR as a novel, automated imaging biomarker for risk stratification in ^82^Rb PET MPI.

**Condensed Abstract:** AI-derived splenic ratio (SR), a marker of pharmacologic stress adequacy, was independently associated with increased cardiovascular risk in a large ^82^Rb PET cohort, even among patients with preserved myocardial flow reserve (MFR). High SR identified individuals with elevated MACE risk despite normal perfusion and flow findings, suggesting unrecognized physiologic vulnerability. Incorporating automated SR into PET MPI interpretation may enhance risk stratification and identify patients who could benefit from intensified preventive care, particularly when traditional imaging markers appear reassuring. These findings support SR as a clinically meaningful, easily integrated biomarker in stress PET imaging.

## Introduction

Rubidium ^82^Rb positron emission tomography (PET) myocardial perfusion imaging (MPI) is increasingly utilized in the evaluation of coronary artery disease (CAD) due to its ability to quantify myocardial blood flow (MBF) and myocardial flow reserve (MFR)(1,2). Compared to traditional single-photon emission computed tomography (SPECT), ^82^Rb PET offers superior image quality making it a valuable tool for both diagnosis and risk stratification (3).

The diagnostic performance of ^82^Rb PET MPI, however, is dependent not only on technical image quality but also on the adequacy of pharmacologic stress. Inadequate stress can reduce test sensitivity for detecting ischemia and increase the likelihood of false-negative results(3,4). Patient-specific factors—such as recent caffeine consumption, underlying autonomic tone, or comorbidities that affect vasodilator response—can attenuate the hyperemic effect of agents like regadenoson. This physiologic variability highlights a limitation of current stress testing protocols and contributes to uncertainty in interpreting visually normal imaging results.

The spleen, as part of the splanchnic circulation, normally exhibits reduced perfusion during pharmacologic stress, a physiologic shift that can be visualized as decreased tracer activity (5,6). This phenomenon, termed splenic switch-off (SSO), reflects appropriate systemic vascular response to stress and has gained attention as a surrogate marker of stress adequacy. The absence of SSO is associated with inadequate stress and potentially diminishes diagnostic accuracy of the stress MPI study (7,8). Additionally, factors such as recent caffeine intake may blunt the splenic response, potentially leading to a false-negative impression of abnormal SSO.

Prior studies have sought to quantify this response using spleen-to-liver-based ratios, showing potential prognostic value (9,10), using liver-normalized spleen-to-spleen uptake comparisons at stress and rest. However, these methods are limited by the need for manual calculation and reader variability, restricting their clinical utility.

In this study, we developed and applied a fully automated, AI-based method to assess splenic response using ^82^Rb PET MPI in a large, multicenter cohort. We examined the relationship between splenic response, MFR, ischemic burden, and clinical outcomes, with the goal of evaluating whether automated analysis of this routinely acquired signal could serve as an adjunctive tool for prognostic stratification and stress adequacy in clinical practice.

## Methods

### Patient Population

A total of 11,795 patients who underwent clinically indicated regadenoson stress PET MPI were drawn from three centers—Cedars-Sinai Medical Center (CSMC), Brigham and Women’s Hospital (BWH), and Intermountain Medical Center—all participating sites in the Registry of Fast Myocardial Perfusion Imaging with Next Generation PET (REFINE PET). The imaging tracer used was ^82^Rb for patients undergoing imaging with regadenoson stress. Patients were excluded who had no spleen or liver visible in the field of view or there was no uptake identified (n=882). In total, 10,913 cases with available stress and rest scans for spleen and liver were included in the analysis: CSMC (n=3482), BWH (n=1590), and Intermountain Medical Center (n=5841). The study protocol received approval from the institutional review boards of all participating institutions. Oversight of the multicenter study was coordinated through the Cedars-Sinai Medical Center IRB. Each site obtained either written informed consent from participants or a waiver of consent for use of de-identified data, as per local regulations.

### Clinical Variables

In this retrospective study, data were collected on participants’ demographic and clinical characteristics, including age, sex, body mass index, and family history of coronary artery disease (CAD). Smoking status and comorbid conditions such as hypertension, dyslipidemia, diabetes, and peripheral artery disease were recorded. Clinical history also included the presence of CAD defined as a history of prior myocardial infarction, previous percutaneous coronary intervention or stents, and previous coronary artery bypass graft surgery. Resting blood pressure and heart rate measurements were obtained before pharmacologic stress testing. Peak stress heart rate and blood pressure, along with clinical and electrocardiogram responses, were collected at the time of clinical reporting, with no distinction made between pharmacologic stress.

Patients were stratified by splenic response and ischemia severity and were categorized into four ischemia severity groups based on their summed difference score (SDS which is defined as the difference between SSS and SRS (based on SSS and SRS from the 17-segment American Heart Association model) (11) . The cutoffs for SDS were defined as: SDS<2 (no ischemia), SDS 2–4 (mild), SDS 5–8 (moderate), and SDS >8 (severe), consistent with prior studies (12).

### PET Imaging and Quantification

Patients underwent rest and stress PET MPI in accordance with clinical guidelines (13). Patients were imaged with ^82^Rb. Early dynamic acquisitions were utilized to quantify stress and rest MBF, with (MFR) calculated as stress myocardial blood flow (MBF) / rest MBF (14,15). Abnormal MFR was defined as <2 (16). The analysis was performed with automated software at the core laboratory with dedicated software (QPET, Cedars-Sinai Medical Center, Los Angeles, California) (15,17).

### CTAC Image Acquisition

Computed Tomography Attenuation Correction (CTAC) scans were acquired according to site-specific protocols. At Brigham and Women’s Hospital, CTAC scans were acquired with tube voltage 120-140 kVp, tube current 10-26 mAs, and slice thickness of 2.5-5 mm. At Cedars-Sinai Medical Center, CTAC scans were acquired with tube voltage 100kVp, tube current 11-13 mAs, and slice thickness of 3 mm. At Intermountain Medical Center, CTAC scans were acquired with tube voltage of 120-130 kVp, tube current of 15-38 mAs, and slice thickness of 2 mm.

Rest and pharmacological stress PET-MPI studies using ^82^Rb were conducted with a Siemens Biograph 64 or Siemens Biograph 1094 (Siemens Healthcare, Erlangen, Germany) for Cedars-Sinai. Brigham and Women’s hospital used a GE Discovery MI, GE Discovery RX, or GE Discovery STE (GE Healthcare, Waukesha, Wisconsin). Intermountain Medical Center used a Siemens Biograph 16, Siemens Biograph 20, or Siemens Biograph 40 (Siemens Healthcare, Erlangen, Germany). A weight-adjusted dose of 609–2424 MBq (16-66 mCi) of ^82^Rb was administered, with a 6-minute rest list-mode acquisition beginning immediately before the injection. Pharmacological stress was induced with regadenoson, and a 6-minute stress list-mode acquisition started concurrently with the infusion of ^82^Rb at the same or similar dosage as the rest scan. As previously described, a low-dose CT scan was performed before each rest and stress PET acquisition to enable attenuation correction (18).

### CTAC Segmentation and SSO Quantification

An artificial intelligence (AI) model was utilized to segment liver and spleen from CTAC scans. These segmentations were performed using the publicly available Total Segmentator model (19). The model utilizes a no new-net (nnU-Net) architecture to automatically segment a variety of anatomic structures from images (20). The nnU-Net automatically adapts to training cases and automatically configures a matching U-Net-based segmentation pipeline. To further the potential utility of applying CT-based segmentation to PET quantification, tracer uptakes were quantified in the liver and spleen volumes during steady state (**Figure 1**).

**Figure 1:**
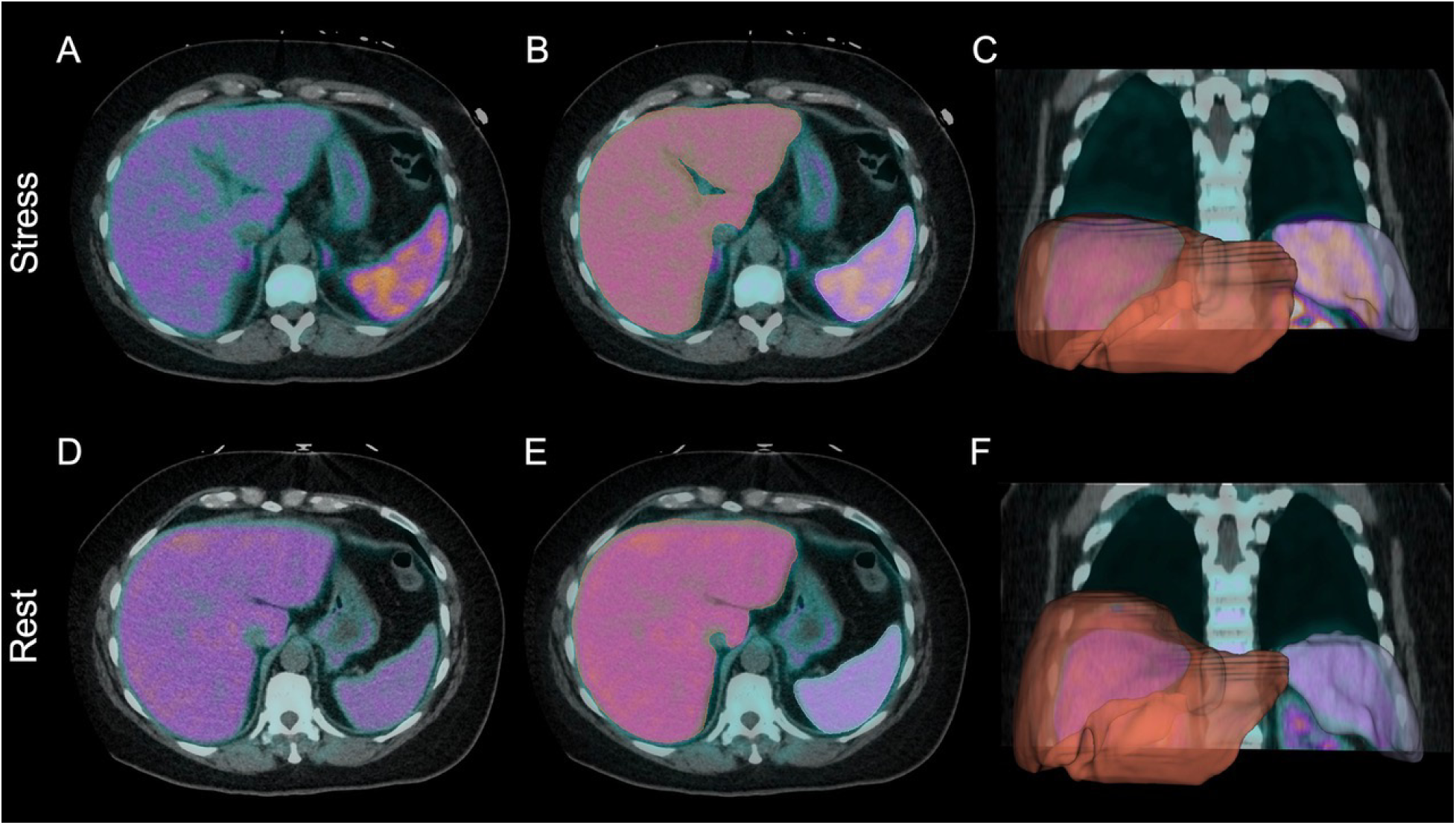
AI-Based Segmentation: Panels A–C show axial and coronal views during stress imaging, while panels D–F display corresponding rest images. PET/CT fusion images (19) illustrate tracer distribution (A, D), with overlaid liver and spleen contours generated by the deep learning nnU-Net AI model (20) (B, E). Three-dimensional reconstructions (C, F) demonstrate organ segmentation accuracy used to derive splenic ratio (SR) by comparing stress and rest spleen activity.

To quantify SSO, splenic ratio (SR) was calculated using the following formula:

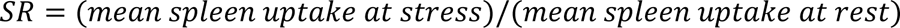

Ratios were divided into deciles, and the cut-point for risk stratification was defined as the lowest decile at which the hazard ratio for adverse outcomes reached statistical significance. Individuals at or above this threshold were classified as having a high splenic ratio, while those below were classified as having a low splenic ratio. The decile process was also repeated with the liver ratio (LR) and splenic response ratio (SRR) to identify thresholds for a high and low LR, as well as a high and low SRR, respectfully (**Supplemental Methods**).

### Outcome

Information on cardiovascular risk factors and clinical outcomes was obtained from review of electronic health care records. The primary outcome was major adverse cardiovascular events (MACE) included all-cause mortality, late revascularization (>90 days after PET MPI scan), myocardial infarction, or admission for unstable angina.

### Statistical Analysis

Continuous variables that were not normally distributed were summarized as median with interquartile range (IQR) and compared using the Wilcoxon rank sum test. Categorical variables were summarized as counts and percentages, and comparisons between groups were performed using Pearson’s chi-squared test. A two-tailed p-value of less than 0.05 was considered statistically significant.

To evaluate the prognostic significance of ischemia severity, 95% confidence interval (CI) error bars were generated using bootstrapping for each of the four SDS categories: <2 (no ischemia), 2–4 (mild), 5–8 (moderate), and >8 (severe). Univariable and multivariable Cox proportional hazards models were constructed to assess associations with MACE, and results are presented as hazard ratios (HRs) with corresponding 95% CIs. Model significance was assessed using the likelihood ratio, Wald, and log-rank tests.

Missing data were handled using simple imputation: missing continuous variables were imputed with the median, and missing categorical variables were imputed with the mode (**Supplemental Table 1)**. Multivariable models were adjusted for age, sex, hypertension, dyslipidemia, diabetes mellitus, BMI, log-transformed CAC score (from rest CTAC or stress CTAC if rest was unavailable), SSS, MFR, and stress LVEF.

When comparing MFR between high and low splenic ratio groups, the same adjustment set was used, excluding MFR. In a separate analysis restricted to patients with preserved flow (MFR ≥2) and no ischemia (SDS <2), the same covariates were used, excluding both MFR and SSS.

To evaluate splenic ratio as a continuous predictor, decile bar charts were constructed by dividing the cohort into ten equal groups. Each decile group was compared to the remainder of the population using Cox models adjusted for the same covariates. Each decile’s minimum and maximum values are rounded to the nearest hundredth just for the visual bar graphs. An additional analysis was performed for the top 5% of splenic ratio values (≥1.13), using the same adjustment strategy.

All statistical analyses were performed using Python version 3.11.5 (Python Software Foundation, Wilmington, DE, USA) and R version 4.4.1 (R Foundation for Statistical Computing, Vienna, Austria).

### Independent Data Access and Analysis

ND, GR, AS, and PS had full access to all the data in the study and take responsibility for its integrity and the data analysis.

## Results

### Baseline Characteristics

**Table 1** demonstrates the clinical and demographic characteristics of the patients stratified by high and low SR. Among the 10,913 patients included, the median age was 68 years, with a majority of male (57%) and White (87%) individuals. Cardiovascular risk factors were common, including hypertension (prevalence 84%), dyslipidemia (76%), diabetes (33%), and current smoking status (25%). Among patients with known coronary artery disease, 20% had a history of PCI and 8.8% had a history of CABG. As some individuals underwent both procedures, these categories are not mutually exclusive. The median body mass index was 28.8 kg/m^2^, and nearly one-quarter of patients had died by the time of analysis. Left ventricular ejection fraction at rest and during stress was preserved overall, with a minority showing reduced function (LVEF <40%). The burden of ischemia and scar, as reflected by SSS and SDS, was low. Median MFR was 2.3, and the log-transformed coronary artery calcium score suggested variable atherosclerotic burden across the cohort.

**Table 1.** Baseline Characteristics and Clinical Outcomes Stratified by Splenic Ratio.

| **Characteristic** | **Overall**<br>**N = 10,913** | **Low**<br>**Splenic Ratio**<br>**N = 9,822** | **High**<br>**Splenic Ratio**<br>**N = 1,091** | **p-value** |
| --- | --- | --- | --- | --- |
| Age | 68 (59, 76) | 68 (59, 76) | 68.0 (59, 76) | 0.5 |
| Male | 6,207 (57%) | 5,568 (57%) | 639 (59%) | 0.2 |
| Race |  |  |  | 0.3 |
| **American Indian or**<br>**Alaska Native** | 50 (0.5%) | 43 (0.5%) | 7 (0.7%) |  |
| **Asian** | 269 (2.6%) | 246 (2.7%) | 23 (2.2%) |  |
| **Black or African**<br>**American** | 875 (8.5%) | 775 (8.4%) | 100 (9.7%) |  |
| **Multiracial** | 9 (<0.1%) | 8 (<0.1%) | 1 (<0.1%) |  |
| **Native Hawaiian or**<br>**Other Pacific**<br>**Islander** | 134 (1.3%) | 116 (1.3%) | 18 (1.8%) |  |
| **White** | 8,952 (87%) | 8,075 (87%) | 877 (85%) |  |
| Smoker | 2,680 (25%) | 2,418 (25%) | 262 (24%) | 0.6 |
| Hypertension | 9,043 (84%) | 8,198 (83%) | 942 (86%) | 0.016 |
| Dyslipidemia | 8,223 (76%) | 7,493 (76%) | 827 (76%) | 0.7 |
| Diabetes Mellitus | 3,559 (33%) | 3,151 (32%) | 408 (37%) | <0.001 |
| Family History | 3,979 (38%) | 3,602 (38%) | 377 (35%) | 0.12 |
| BMI (kg/m<sup>2</sup>) | 28.8 (25.1, 33.8) | 28.7 (25.0, 33.5) | 30.8 (26.2, 37.1) | <0.001 |
| Previous CAD | 3,333 (31%) | 2,999 (31%) | 334 (31%) | >0.9 |
| Past MI | 2,073 (19%) | 1,861 (19%) | 212 (19%) | 0.7 |
| Past PCI or stents | 2,165 (20%) | 1,963 (20%) | 202 (19%) | 0.3 |
| Past Bypass surgery | 958 (8.8%) | 878 (8.9%) | 80 (7.3%) | 0.084 |
| Death | 2,655 (24%) | 2,340 (24%) | 315 (29%) | <0.001 |
| Rest LVEF | 65.6 (54.7, 73.4) | 65.9 (54.9, 73.6) | 63.9 (53.0, 71.4) | <0.001 |
| Stress LVEF | 70.0 (58.4, 77.5) | 70.2 (58.8, 77.8) | 67.3 (56.4, 74.6) | <0.001 |
| Rest LVEF < 40% | 1,151 (11%) | 1,027 (10%) | 124 (11%) | 0.4 |
| Positive Stress ECG | 246 (2.3%) | 225 (2.3%) | 21 (1.9%) | 0.5 |
| CAC score | 152.0 (0.0, 967.0) | 153.3 (0.0, 975.7) | 143.3 (0.0, 909.6) | 0.6 |
| MFR | 2.3 (1.8, 2.9) | 2.3 (1.8, 2.9) | 2.2 (1.8, 2.8) | <0.001 |
| SSS | 0 (0, 4) | 0 (0, 4) | 0 (0, 5) | 0.035 |
| SDS | 0 (0, 2) | 0 (0, 2) | 0 (0, 2) | >0.9 |
| MACE | 3444 (32%) | 3,047 (31%) | 397 (36%) | 0.001 |
| All Cause-Mortality | 2655 (24%) | 2,340 (24%) | 315 (29%) | <0.001 |
| Non-Fatal MI | 579 (5.3%) | 504 (5.1%) | 75 (6.9%) | 0.016 |
| Late Revascularization | 682 (6.2%) | 605 (6.2%) | 77 (7.1%) | 0.26 |

Continuous variables are presented as median (interquartile range) and were compared using the Wilcoxon rank sum test. The categorial variables are shown by n (%) and were compared by Pearson's Chi-squared test. Abbreviations: BMI – body mass index; CAD – coronary artery disease; MI – myocardial infarction; PCI – percutaneous coronary artery; LVEF – left ventricular ejection fraction; ECG – electrocardiogram; CAC – coronary artery calcium; MFR –
myocardial flow reserve, SSS – summed stress score; SDS – summed difference score; MACE – mace adverse cardiovascular events

### Determination of Splenic Ratio Thresholds

To determine the optimal SR cutoff for risk stratification, we applied decile-based stratification as shown in **Figure 2, Supplemental Figure 1**, and **Supplemental Figure 2**. The tenth decile (≥1.03) was selected as the threshold, as it marked the point at which the hazard ratio for MACE became significantly elevated. Patients with SR values below this threshold (<1.03) were categorized as having a low splenic ratio, and those at or above the threshold (≥1.03) were classified as having a high splenic ratio, indicating an attenuated splenic response to pharmacologic stress.

**Figure 2.**
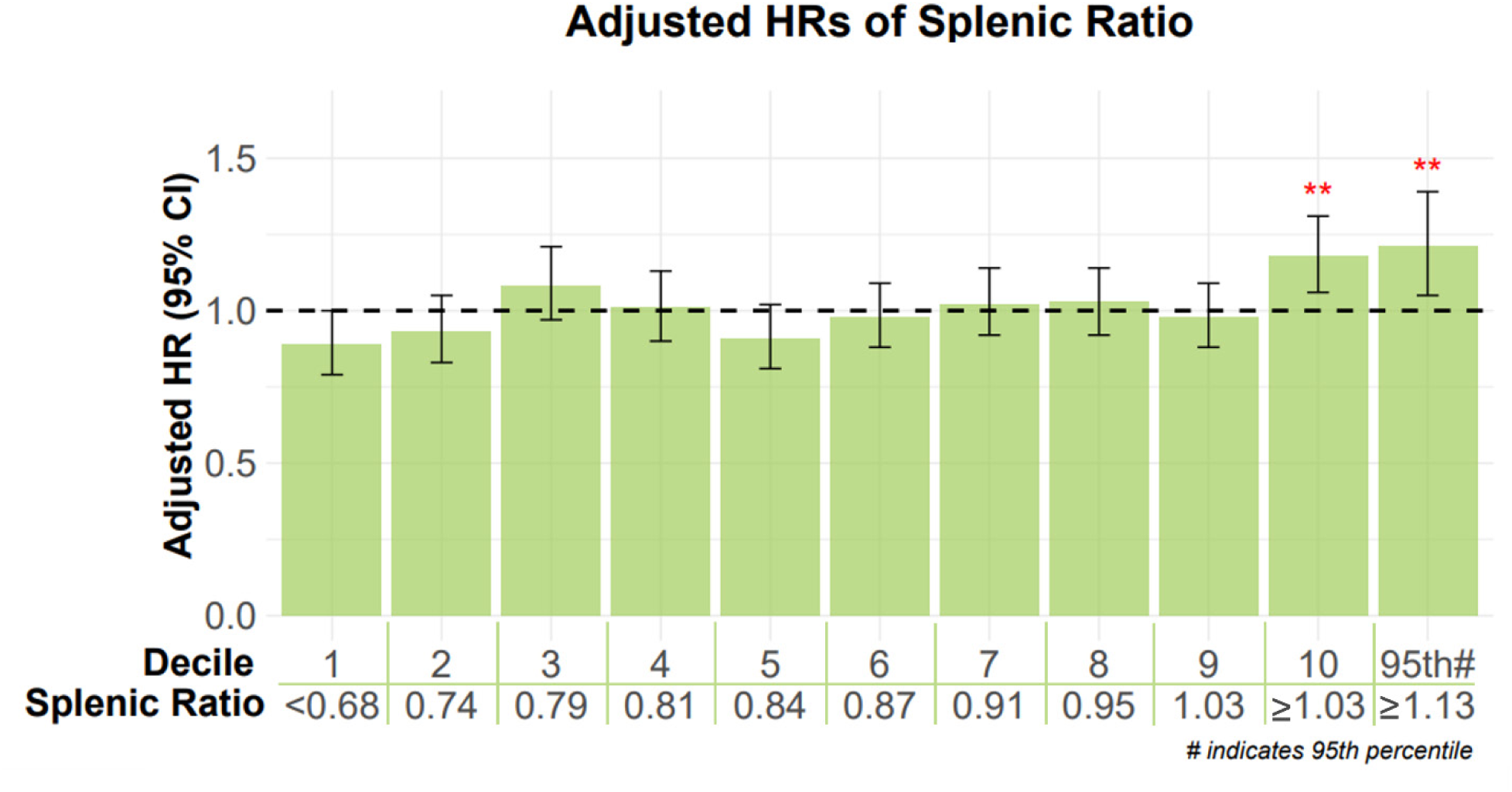
Adjusted HRs of Splenic Ratio: Adjusted hazard ratios with 95% CIs for MACE outcomes across deciles of the stress/rest splenic ratio. Each bar represents a decile group marked from 1 to 10 or the 95^th^ percentile of the splenic ratio, with the rest of the population used as the reference group. The x-axis shows the decile rank (or 95^th^ percentile) as the first row with the maximum splenic ratio value in the second row from decile 1 to 9. Decile 10 and the 95^th^ percentile hold the minimum splenic ratio value which is 1.03 and 1.13 respectively. The red asterisks show the statistical significance for decile 10 and 95^th^ percentile with p<0.01. Abbreviations: HR (Hazard Ratio), CI (Confidence Interval)

### Risk of MACE Across Splenic Ratio Deciles

Figure 2 shows the adjusted HRs of MACE across the splenic ratio deciles for the entire study population. As shown in the figure, only the 10^th^ decile and the 95^th^ percentile have a strong statistical significance of p<0.01, meaning patients with a splenic ratio greater than or equal to 1.03 have a significant increased risk of MACE compared to patients with less than a 1.03 splenic ratio. This is further statistically significant of p<0.001 for patients with MFR≥2 at the 10^th^ decile and 95^th^ percentile. Even the 1^st^ decile was statistically significant of p<0.01, meaning patients below 0.68 are less likely to experience a MACE event than patients above this threshold. However, for patients with MFR<2, there is no statistical significance at the cutoff 1.03 or at any decile for the adjusted hazard ratios (**Supplemental** Figure 2). Detailed adjusted HRs for SRR and LR are shown in **Supplemental** Figure 3 **and Supplemental** Figure 4 **respectively.**

### Clinical Outcomes of Splenic Response

Patients with a high SR demonstrated higher prevalence of cardiovascular risk factors compared to those with a low splenic ratio as shown in **Table 1**. The incidence of MACE and ACM was higher in the high splenic ratio group than the low splenic ratio group (36% vs. 31% for MACE; 29% vs. 24% for ACM) respectfully, with both outcomes reaching statistical significance (p=0.001 and p<0.001 respectfully). Non-fatal myocardial infarction was more common in patients with elevated splenic ratio (6.9% in the high SR group vs. 5.1% in the low SR group; p=0.016). Late revascularization occurred in 7.1% of patients with high SR and 6.2% with low SR, though this difference was not statistically significant (p=0.26). In addition, patients with a high splenic ratio had a higher prevalence of diabetes mellitus (37% vs. 32%; p<0.001) and hypertension (86% vs. 83%; p=0.016), and a higher median body mass index (30.8 vs. 28.7 kg/m²; p<0.001).

As shown in **Table 2**, among patients with preserved MFR, those with a high splenic ratio experienced significantly worse clinical outcomes. Rates of major adverse cardiovascular events (29% vs. 22%, p < 0.001), all-cause mortality (22% vs. 16%, p < 0.001), and non-fatal myocardial infarction (5.4% vs. 3.7%, p = 0.045) were all significantly higher in the high SR group compared to the low SR group. In contrast, the rate of late revascularization (5.5% vs. 4.7%, p = 0.36) did not differ significantly between groups.

**Table 2.** Clinical Outcomes by Splenic Ratio: MFR≥2.

| **Characteristic** | **Low Splenic**<br>**Ratio**<br>**N = 6,623** | **High Splenic**<br>**Ratio**<br>**N = 687** | **p-value** |
| --- | --- | --- | --- |
| Major Adverse Cardiovascular Events (MACE) | 1,451 (22%) | 202 (29%) | <0.001 |
| All-Cause Mortality | 1,030 (16%) | 151 (22%) | <0.001 |
| Non-Fatal Myocardial Infarction | 245 (3.7%) | 37 (5.4%) | 0.045 |
| Late Revascularization (PCI/CABG) | 313 (4.7%) | 38 (5.5%) | 0.36 |

Categorical variables are shown by n (%) and were compared by Pearson's Chi-squared test.
Abbreviations: PCI – percutaneous coronary artery; CABG – coronary artery bypass grafting

In the overall cohort, the five-year incidence of MACE in Figure 3A was higher in patients with a high SR compared to those with a low SR, particularly in those with no ischemia (SDS <2), (26% [n=767] vs. 21% [n=6938], p=0.0004). No significant differences were observed in the mild, moderate, or severe ischemia groups. In contrast, among patients with preserved MFR (MFR≥2) in Figure 3B, the adverse impact of a high splenic ratio was more pronounced.

**Figure 3.**
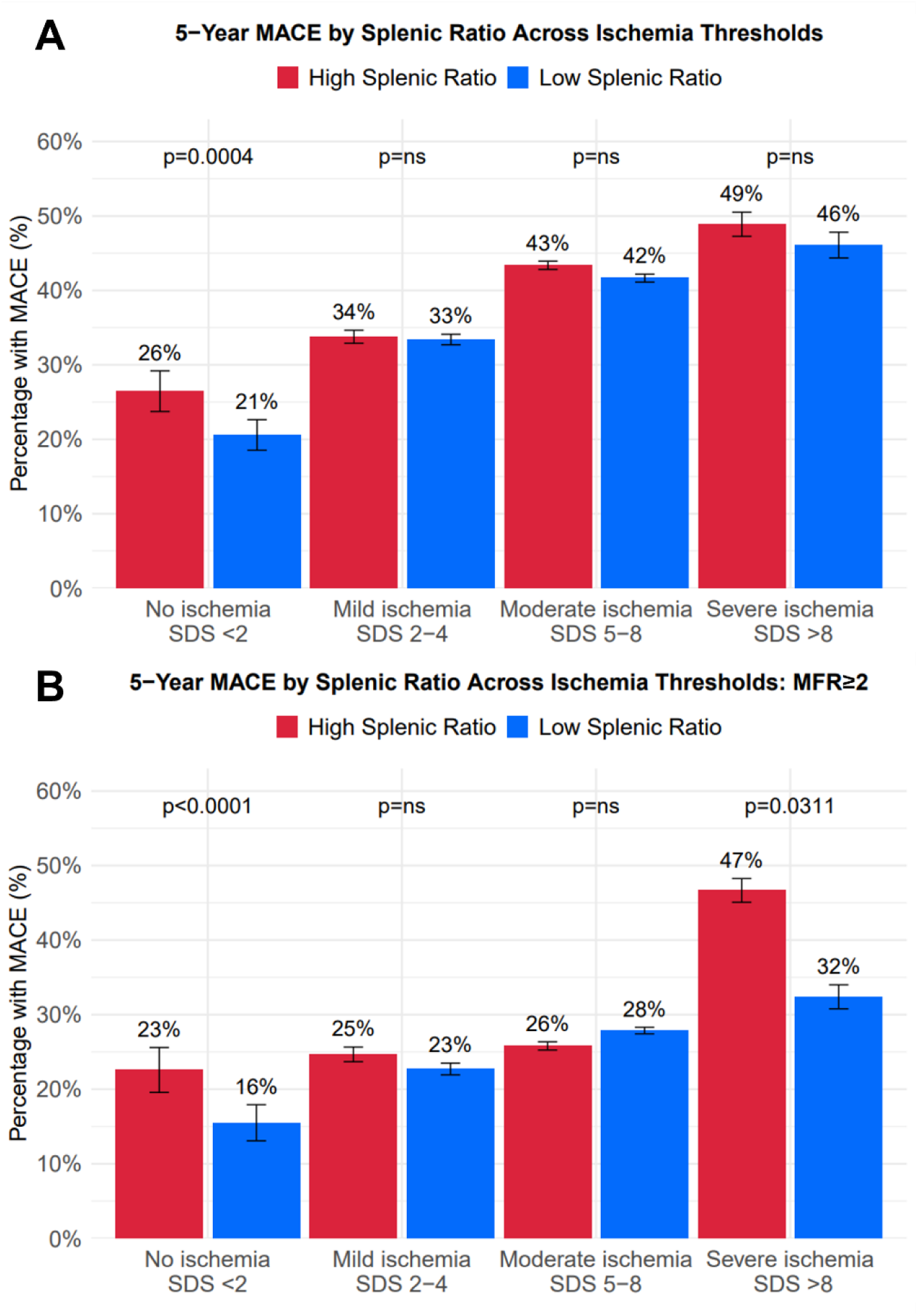
Five-Year MACE Rates by Splenic Ratio Across Ischemic Categories, Stratified by Ischemia Thresholds. Five-year MACE rates by splenic ratio (SR) across ischemic categories in the overall cohort (A) and among patients with preserved myocardial flow reserve (MFR ≥2) (B). If p-value ≥0.05, p=ns is noted.

Significant differences in MACE were observed in both the no ischemia (23% [n=549] vs. 16% [n=5256], p<0.0001) and severe ischemia (47% [n=30] vs. 32% [n=213], p=0.0311) subgroups (Figure 3B). However, the statistical significance observed in the severe ischemia subgroup should be interpreted with caution due to the small sample size.

In multivariable Cox regression analysis adjusting for key clinical, metabolic, and imaging covariates, a high splenic ratio (10th decile) was independently associated with an increased risk of MACE in both the overall cohort and the subgroup with preserved MFR (MFR ≥2). In the full cohort (Figure 4A), high splenic ratio was associated with a hazard ratio (HR) of 1.18 (95% CI: 1.06–1.31; p=0.002). In the MFR ≥2 subgroup (Figure 4B), the association was even stronger, with an HR of 1.44 (95% CI: 1.24–1.66; p<0.001), representing one of the highest effect sizes among all covariates. Traditional cardiovascular risk factors—such as hypertension, diabetes mellitus, and elevated CAC score—also demonstrated significant associations with adverse outcomes in the overall cohort.

**Figure 4.**
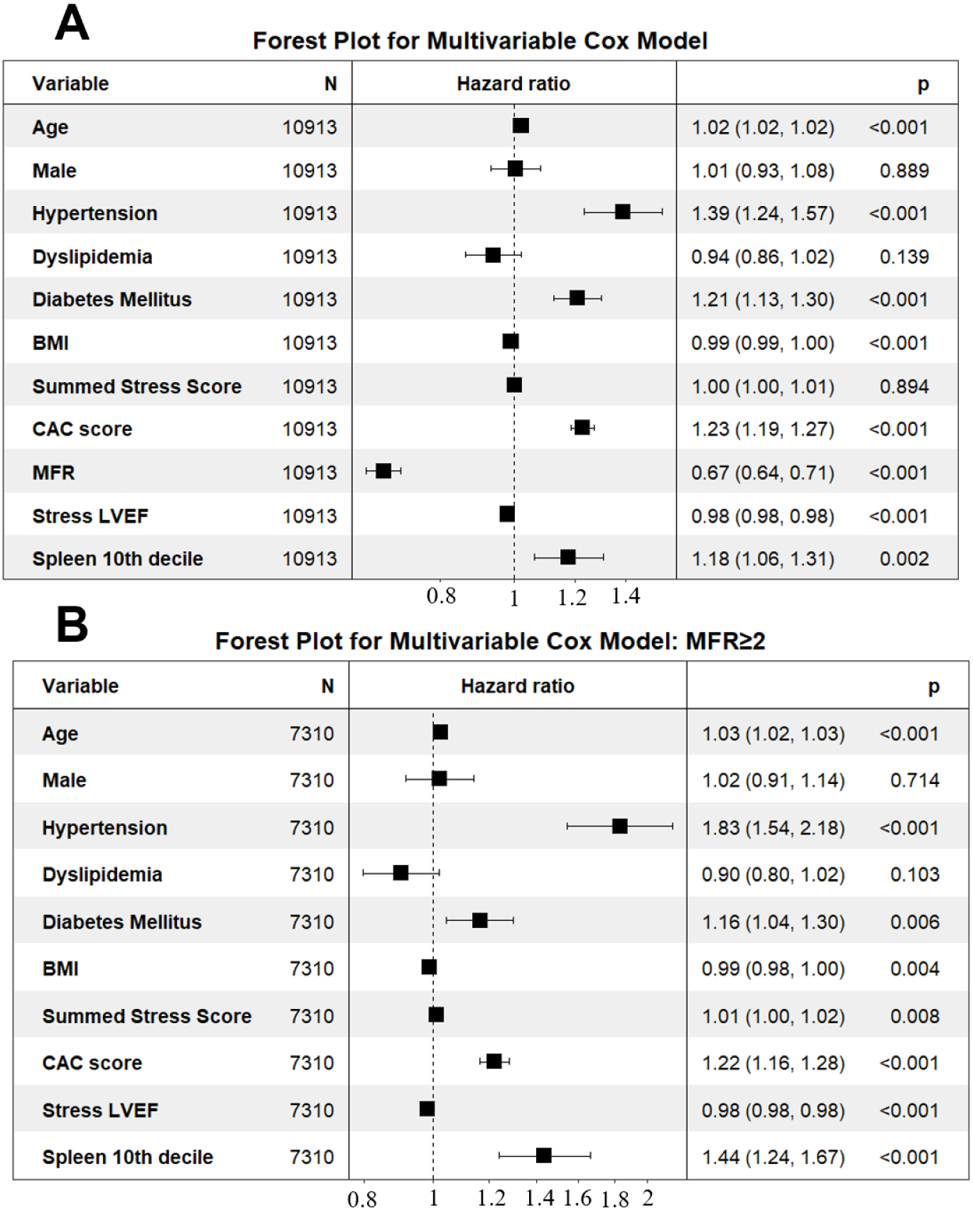
Multivariable Cox Regression for MACE: Association of High Splenic Ratio (10th Decile) with Outcomes in the Full Cohort and in Patients with MFR ≥2. Multivariable Cox regression for MACE showing the association of high splenic ratio (≥90th percentile) with outcomes in the full cohort (A) and in patients with preserved myocardial flow reserve (MFR ≥2) (B). High splenic ratio remained an independent predictor of adverse events after adjustment for clinical and imaging covariates, with a stronger effect size observed in the preserved MFR subgroup (HR 1.44, 95% CI 1.24-1.67, p < 0.001). These findings highlight SR as a robust, additive biomarker of cardiovascular risk beyond traditional predictors.

In the overall cohort, patients with a high splenic ratio had a higher incidence of MACE compared to those with a low splenic ratio (unadjusted HR 1.20, 95% CI 1.08–1.33, p=0.00059; adjusted HR 1.18, 95% CI 1.06–1.31; Figure 5A). Among patients with preserved MFR (MFR ≥2; n = 7,310), high splenic ratio was associated with significantly lower MACE-free survival (unadjusted HR 1.41, 95% CI 1.22–1.64, p< 0.0001; adjusted HR 1.44, 95% CI 1.24–1.67; Figure 5C). In contrast, among patients with MFR <2 (n = 3,603), there was no statistically significant difference in MACE incidence between high and low splenic ratio groups (unadjusted HR 0.93, 95% CI 0.80–1.08, p = 0.33; adjusted HR 0.99, 95% CI 0.85–1.15; Figure 5D).

**Figure 5.**
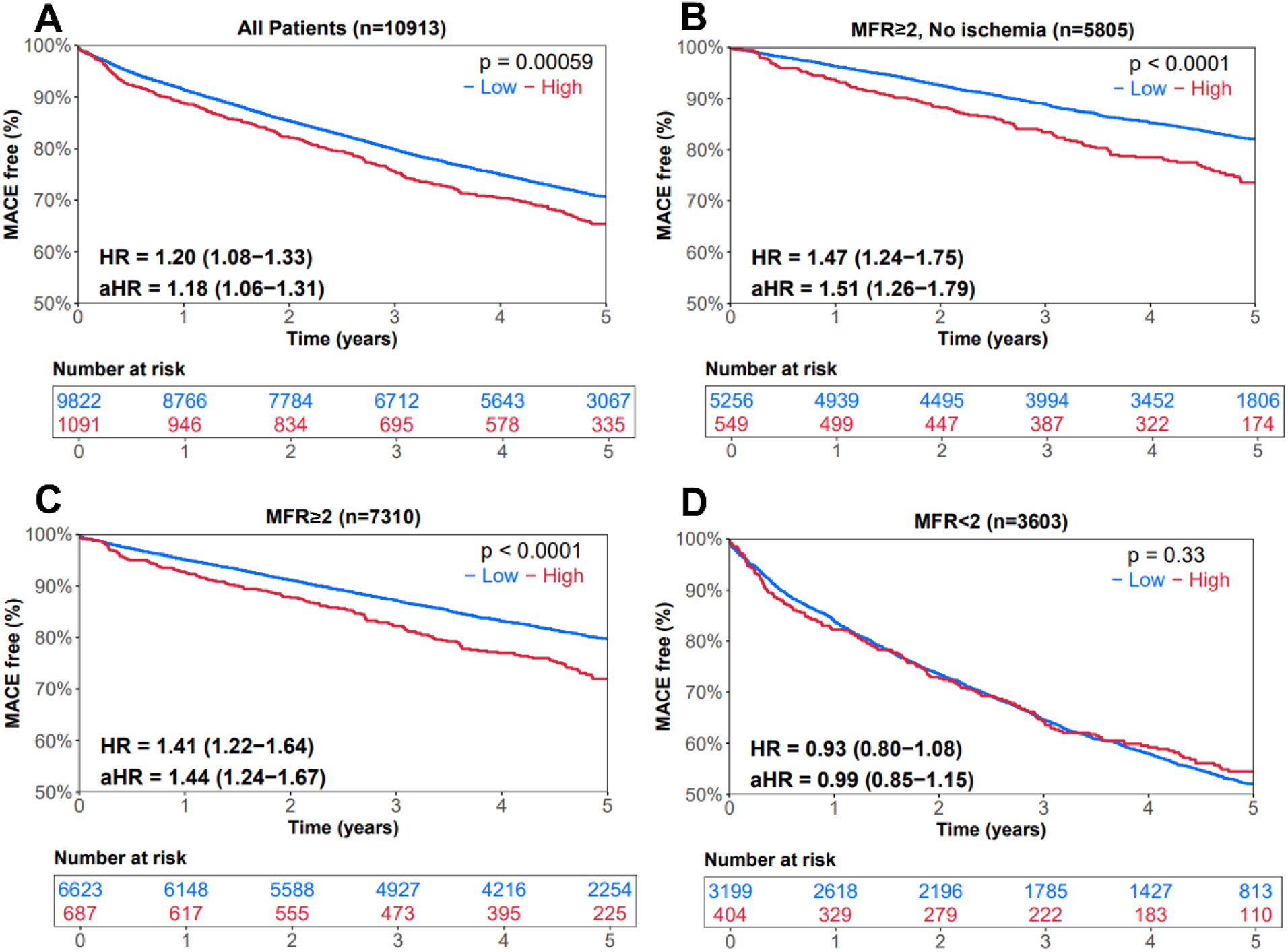
Kaplan–Meier Analysis of 5-Year MACE by Splenic Ratio and MFR. Kaplan– Meier analysis of 5-year MACE by splenic ratio (SR) and myocardial flow reserve (MFR). (A) High SR was associated with significantly lower MACE-free survival in the full cohort. (B–C) This association was stronger in patients with preserved MFR, especially in those without ischemia (SDS <2), with adjusted HRs exceeding 1.41. (D) In contrast, SR did not differentiate outcomes in patients with impaired MFR (<2), suggesting SR adds prognostic value primarily when perfusion and flow appear preserved. Abbreviations: aHR – adjusted hazard ratio (95% confidence interval); HR – unadjusted hazard ratio (95% confidence interval); MACE – Major Adverse Cardiovascular Events; MFR – myocardial flow reserve

These findings were further supported by Kaplan–Meier analyses stratified by both SR and MFR categories, as shown in **Supplemental** Figure 5. Event-free survival was highest in the low SR/preserved MFR group and progressively decreased across the remaining strata, with the lowest survival observed in patients with both high SR and reduced MFR. Adjusted hazard ratios for MACE were 1.41 (95% CI 1.22–1.64) for high SR/MFR ≥2, 1.79 (95% CI 1.66–1.94) for low SR/MFR <2, and 1.80 (95% CI 1.55–2.10) for high SR/MFR <2, all compared to the reference group (low SR/MFR ≥2), further emphasizing the additive prognostic impact of elevated SR and impaired flow.

In the subgroup with both no ischemia (SDS <2) and preserved MFR (n = 5,805) in Figure 5B, high splenic ratio was associated with a significantly higher incidence of MACE over five years (unadjusted HR 1.47, 95% CI 1.24–1.75, p<0.0001; adjusted HR 1.51, 95% CI 1.26–1.79), despite these patients being traditionally considered low-risk based on perfusion and flow measures. The number of patients at risk at each time point is shown beneath each Kaplan–Meier curve.

## Discussion

To the best of our knowledge, this study represents the first comprehensive multicenter evaluation of splenic ratio using a fully automated, deep learning-based approach in patients undergoing ^82^Rb PET regadenoson stress testing and first report of findings in patients with preserved MFR. By leveraging AI algorithms, we eliminated observer variability, which enables integration of SR assessment into clinical workflows. Our findings from a substantially larger cohort compared to prior studies demonstrate that a high SR is independently associated with increased risks of MACE, all-cause mortality, and myocardial infarction—even after adjustment for clinical risk factors, coronary calcification, ischemic burden (SDS), and MFR—and importantly, identifies elevated mortality risk among patients with preserved MFR.

These new insights suggest that SR and MFR represent distinct but complementary physiologic measures. Whereas MFR assesses primarily coronary-specific microvascular health(21), SR may integrate systemic autonomic, vascular, or inflammatory mechanisms. High SR thus appears to reflect broader systemic physiologic vulnerability, explaining its prognostic impact in patients with normal or preserved myocardial perfusion and MFR.

The physiologic mechanisms underlying the observed relationship between SR and cardiovascular outcomes likely reflect broader systemic responses to pharmacologic stress. While traditionally, splenic vasoconstriction has been considered primarily a marker of adequate pharmacologic stress, our findings suggest a more complex physiologic integration involving autonomic regulation, vascular tone, and systemic inflammation (22,23). Impaired autonomic responsiveness and splanchnic vascular dysregulation could contribute to attenuated splenic vasoconstriction during stress(24),(25). Additionally, the spleen plays a role in neuroimmune signaling and acts as a reservoir of inflammatory cells; altered splenic perfusion patterns may thus reflect subclinical endothelial dysfunction or inflammatory states, both independently associated with adverse cardiovascular events(26). These pathways may explain why high SR identifies heightened cardiovascular risk, particularly among patients with preserved myocardial perfusion and flow reserve.

Further analysis stratified by SDS reinforced the prognostic utility of the splenic ratio. As shown in Figure 3A, among patients with no visual ischemia (SDS <2), those with high SR had a significantly higher five-year MACE rate compared to those with low SR. These associations were more pronounced in patients with preserved MFR ≥2 (Figure 3B), particularly within the SDS <2 subgroup, where the five-year MACE rate was higher SR group compared to low SR group. This highlights SR’s potential to unmask residual cardiovascular vulnerability not captured by conventional perfusion or flow metrics.

The prognostic value of SR also varied meaningfully by MFR status. While patients with high SR had a higher prevalence of impaired MFR, the impact of SR on outcomes was largely restricted to those with preserved MFR. Among patients with impaired MFR, SR did not offer additional prognostic stratification. Together, these findings suggest that SR and MFR represent complementary but distinct dimensions of cardiovascular risk. SR may reflect systemic or autonomic influences—such as inflammatory activation or altered sympathetic signaling—not fully captured by MFR alone.

Our multivariable Cox analysis further underscored the independent prognostic value of high SR. Adjusting for traditional cardiovascular risk factors, coronary calcium scores, ischemia severity, left ventricular function, and MFR, high SR remained significantly associated with increased risk for adverse cardiovascular outcomes. The persistence of SR’s prognostic significance in multivariable models highlights its value as an independent marker, distinct from and additive to established predictors.

Clinically, integrating automated SR assessment into PET MPI interpretation could substantially improve patient risk stratification. Patients identified by SR as high-risk, despite preserved MFR or minimal ischemia, may benefit from more aggressive preventative strategies. This highlights SR’s potential role as an adjunctive tool for personalized cardiovascular risk assessment. Prospective studies are warranted to validate these findings further, and to explore whether management approaches informed by SR could lead to improved clinical outcomes, particularly among those not identified by traditional imaging metrics alone.

The strengths of our study include its large, multicenter cohort; the use of fully automated, standardized AI-based SR measurements to reduce operator variability; and the assessment of prognostic value across a range of perfusion and flow parameters, including in patients with preserved MFR. The automated nature of the AI algorithm enhances efficiency and consistency in SR calculation, eliminates observer variability, and offers an approach for efficient clinical interpretation. Our results also suggest that SR is a valuable prognostic tool beyond simply identifying inadequate stress response. A reduced splenic response is associated with a greater ischemic burden, as indicated by higher SSS and lower MFR, pointing to a potential role for SR in guiding clinical management and therapeutic decisions.

Our study has limitations. We rely on the registration of PET and CT to quantify splenic ratio which was performed during clinical quality controls. Any remaining PET/CT misregistration can affect the accuracy of the SR derivation. While our results support the utility of SR in predicting adverse outcomes, prospective validation is necessary to further confirm its role in guiding clinical decision-making. Future research should also assess the potential integration of SR into existing CAD risk models and explore the clinical outcomes associated with SR-based interventions.

## Conclusion

Our multicenter study demonstrates that automated AI-derived SR provides independent prognostic value in patients undergoing ^82^Rb PET MPI, in patients with preserved MFR extending risk stratification beyond conventional MPI assessment. These findings reinforce the feasibility and clinical utility of integrating automated SR into clinical practice and can provide additional physiologic insights into systemic vascular and autonomic responses captured by splenic perfusion patterns.

### Clinical Perspectives

#### Competency in Medical Knowledge

Splenic ratio (SR), derived from ^82^Rb PET myocardial perfusion imaging, reflects the systemic physiologic response to vasodilator stress. High SR is associated with increased cardiovascular risk, particularly among patients with preserved myocardial flow reserve (MFR), highlighting the potential role of SR as an adjunctive prognostic marker.

#### Competency in Patient Care

Patients undergoing PET MPI with preserved MFR and no ischemia are traditionally considered low risk. However, a high SR may indicate residual systemic vulnerability. Recognizing this can prompt clinicians to consider more aggressive preventive strategies even in patients with otherwise reassuring imaging findings.

#### Translational Outlook 1

The findings suggest that SR offers complementary information to MFR and perfusion, capturing systemic or autonomic dysfunction not readily apparent with conventional markers. Future prospective studies should evaluate whether incorporating SR into risk models improves prediction of major adverse cardiovascular events and whether SR-guided preventive strategies improve outcomes in patients with preserved MFR.

#### Translational Outlook 2

Automated AI-based quantification of SR from routinely acquired PET/CT scans offers a scalable and reproducible tool for risk stratification. Implementation studies should assess whether real-time integration of SR into clinical PET MPI workflows enhances diagnostic confidence and facilitates earlier identification of high-risk individuals otherwise missed by standard interpretations.

## Abbreviations List

PET: Position Emission Tomography SSO: Splenic Switch-off
MFR: Myocardial Flow Reserve
SPECT: Single-Photon Emission Computed Tomography MPI: Myocardial Perfusion Imaging
SSS: Summed Stress Score SDS: Summed Difference Score
CTAC: Computed Tomography Attenuation Correction SR: Splenic Ratio

## Data Availability

To the extent allowed by data sharing agreements and IRB protocols, the deidentified data and data analysis code from this manuscript will be shared upon written request.

---

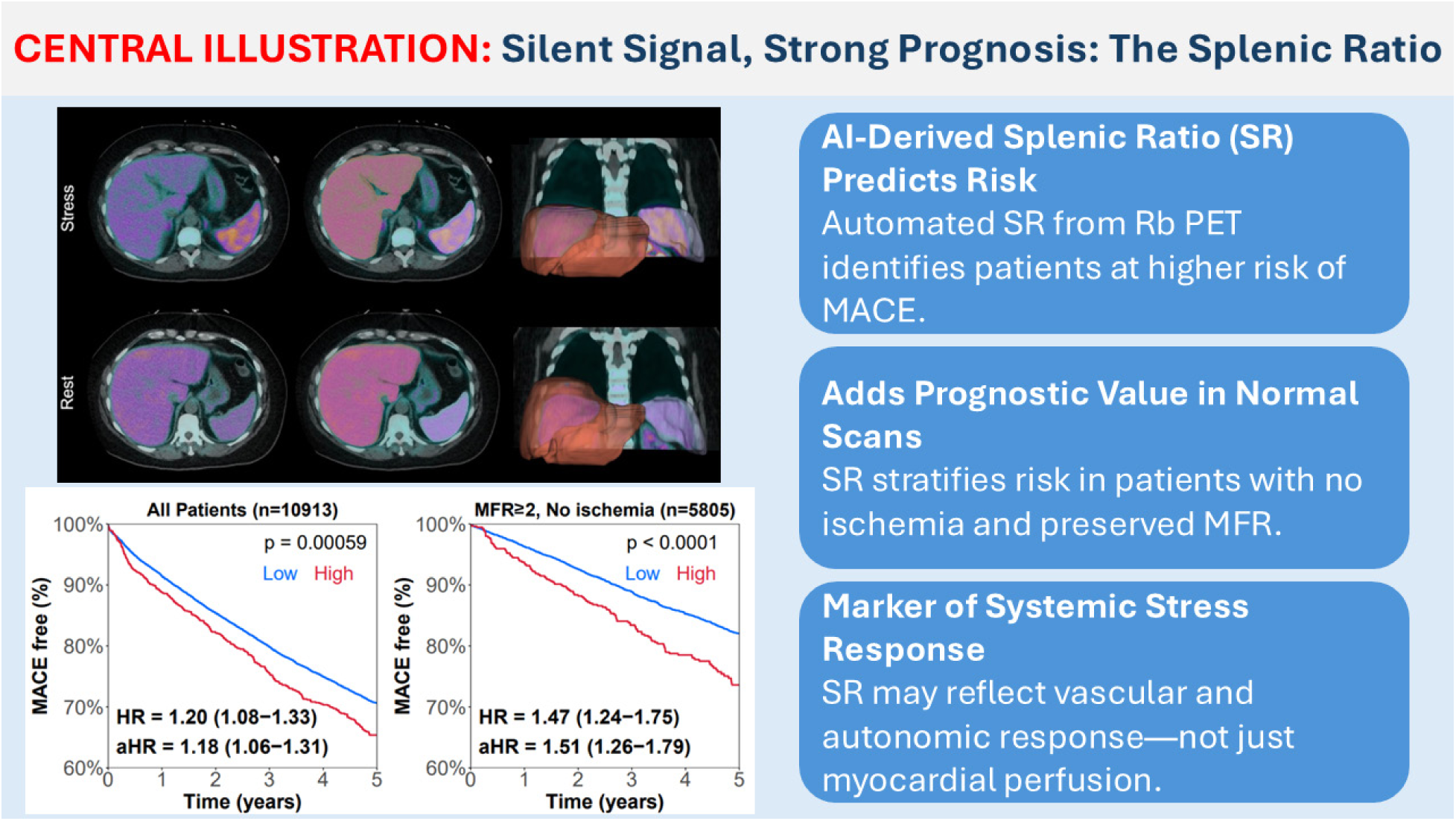
Central Illustration: AI-derived splenic ratio (SR) from ^82^Rb PET imaging identifies increased risk of MACE, particularly in patients with preserved myocardial flow reserve and no ischemia. SR reflects systemic stress response and adds prognostic value beyond traditional perfusion and flow metrics.

