## Supplemental Appendix for "AI-Derived Splenic Response in Cardiac PET Predicts Mortality: A Multi-Site Study"

**Supplemental Material:**

**Supplemental Methods:**

We also replicated the liver-normalized splenic response ratio (SRR) approach described by Bami et al. and Saad et al. (9,10). This method assumes a stable, linear relationship between liver uptake at stress and rest to account for systemic factors. In our large multicenter cohort, liver activity demonstrates a non-linear and bidirectional association with clinical outcomes across its distribution (**Supplemental Figure 4**), raising concerns about potential bias introduced by liver normalization. This variability may have been further amplified by methodological differences, as prior studies sampled only a small region of the liver, while our analysis averaged tracer counts across the entire liver volume within the field of view.

Given these limitations, we elected to simplify the calculation by using spleen stress-to-rest counts alone, defining the splenic ratio (SR) independent of liver uptake. Using decile-based stratification, the highest SR decile consistently identified patients at elevated cardiovascular risk (36% vs. 31% 5-year MACE), validating SR as a robust and clinically meaningful prognostic marker without the need for liver normalization.

$$SRR = \frac{mean\ spleen\ uptake\ at\ stress}{mean\ liver\ uptake\ at\ stress} / \frac{mean\ spleen\ uptake\ at\ rest}{mean\ liver\ uptake\ at\ rest}$$

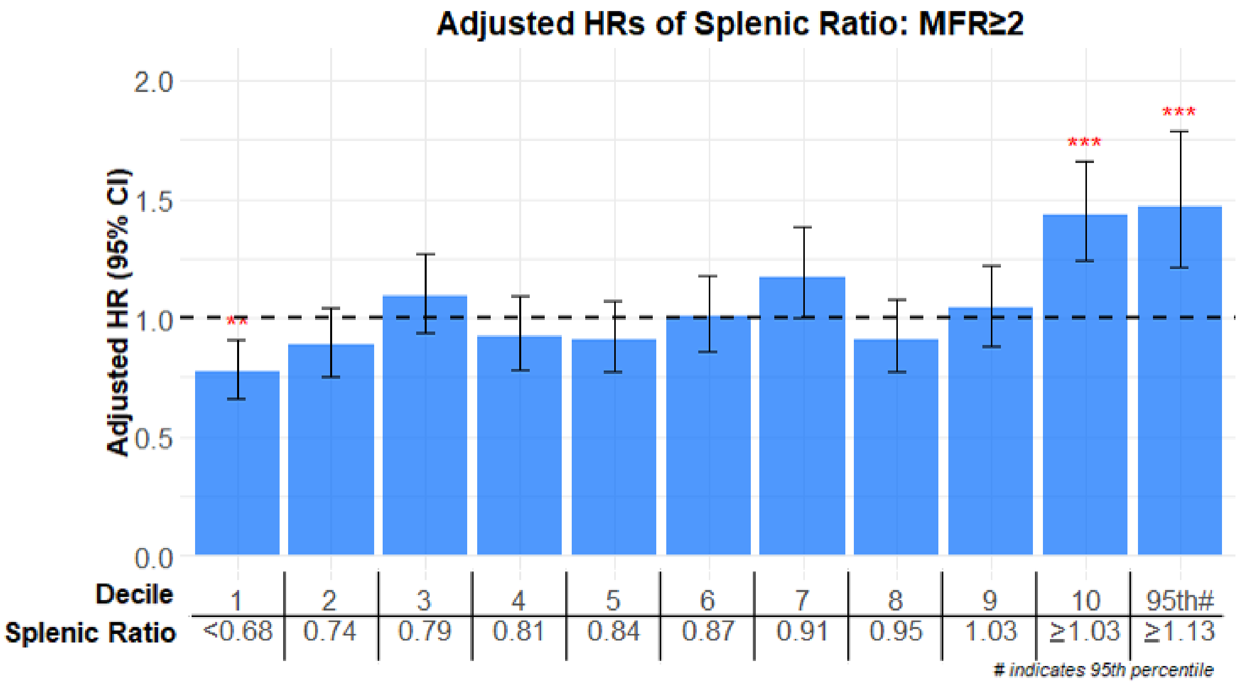


**Supplemental Figure 1. Adjusted hazard ratios for major adverse cardiovascular events (MACE) across deciles of the splenic stress/rest ratio in patients with MFR ≥2 (n=7,310).** Adjusted hazard ratios demonstrated a consistent or significant association between splenic ratio and outcomes in this subgroup, especially at decile 1 and 10 (including the $95^{th}$ percentile). The red asterisks demonstrate the statistical significance compared to the rest of the population. Two asterisks show $p<0.01$ and three asterisks show $p<0.001$.

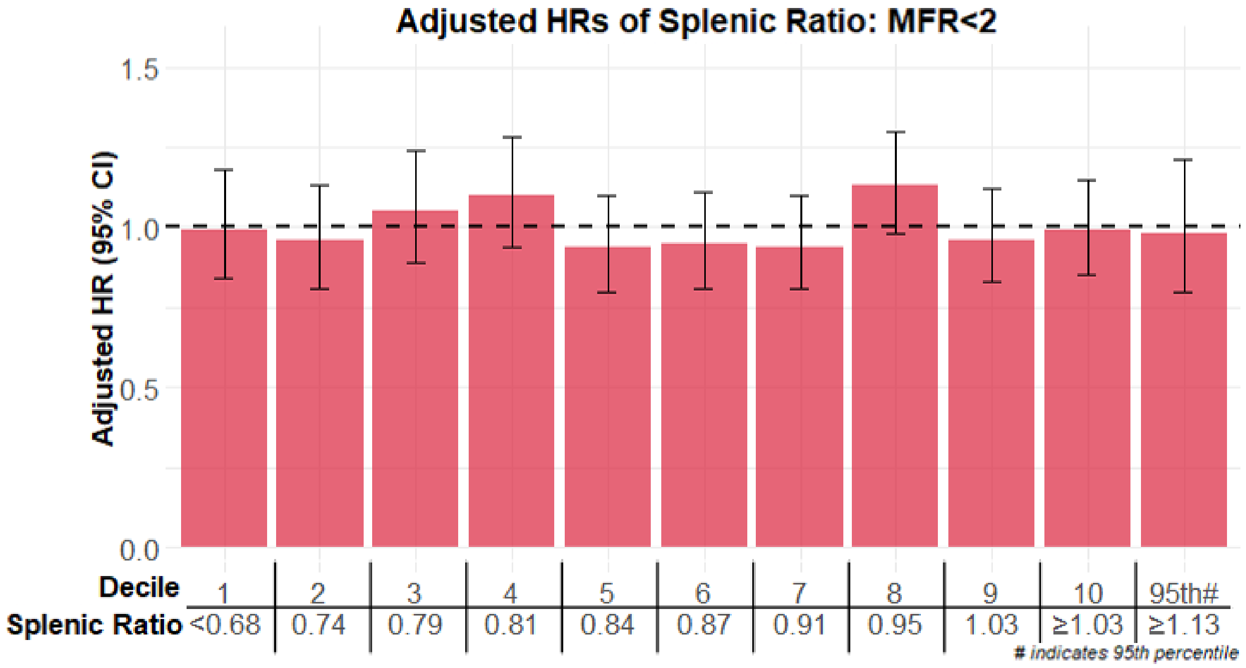


**Supplemental Figure 2. Adjusted hazard ratios for major adverse cardiovascular events (MACE) across deciles of the splenic stress/rest ratio in patients with MFR <2 (n=3,603).** Neither unadjusted nor adjusted hazard ratios demonstrated a consistent or significant association between splenic ratio and outcomes in this subgroup. The bar plot and tabulated hazard ratios confirm the absence of a clear risk gradient across SR percentiles.

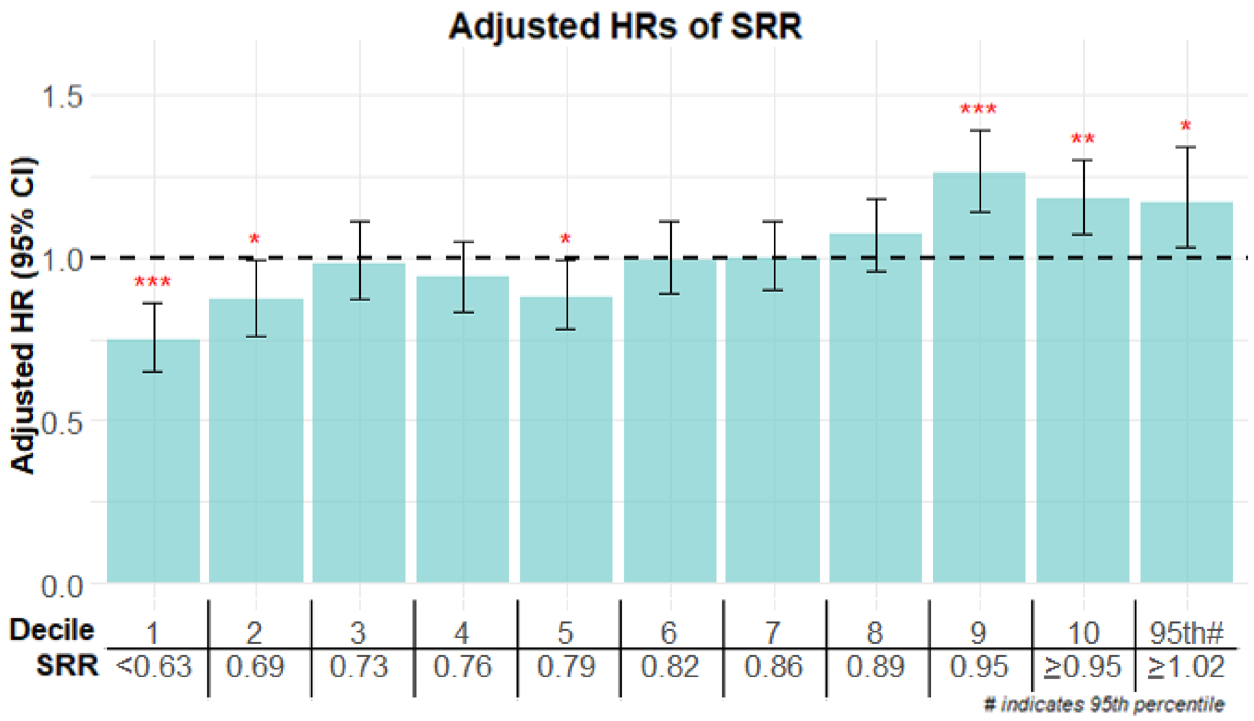


**Supplemental Figure 3. Adjusted hazard ratios for MACE by SRR Percentile Deciles**

Patients were grouped by deciles of the splenic-to-hepatic ratio (SRR) and assigned binary indicators for each range. The plot shows adjusted hazard ratios (HRs) for MACE with 95% confidence intervals, derived from multivariable Cox regression models. Significant deciles ($p < 0.05$) are indicated with red asterisks: *$p < 0.05$, **$p < 0.01$, ***$p < 0.001$. Higher SRR values (≥9th decile) were associated with increased MACE risk, while lower SRR values (1st and 2nd deciles) were associated with reduced risk. The 95th percentile threshold is noted on the far right.

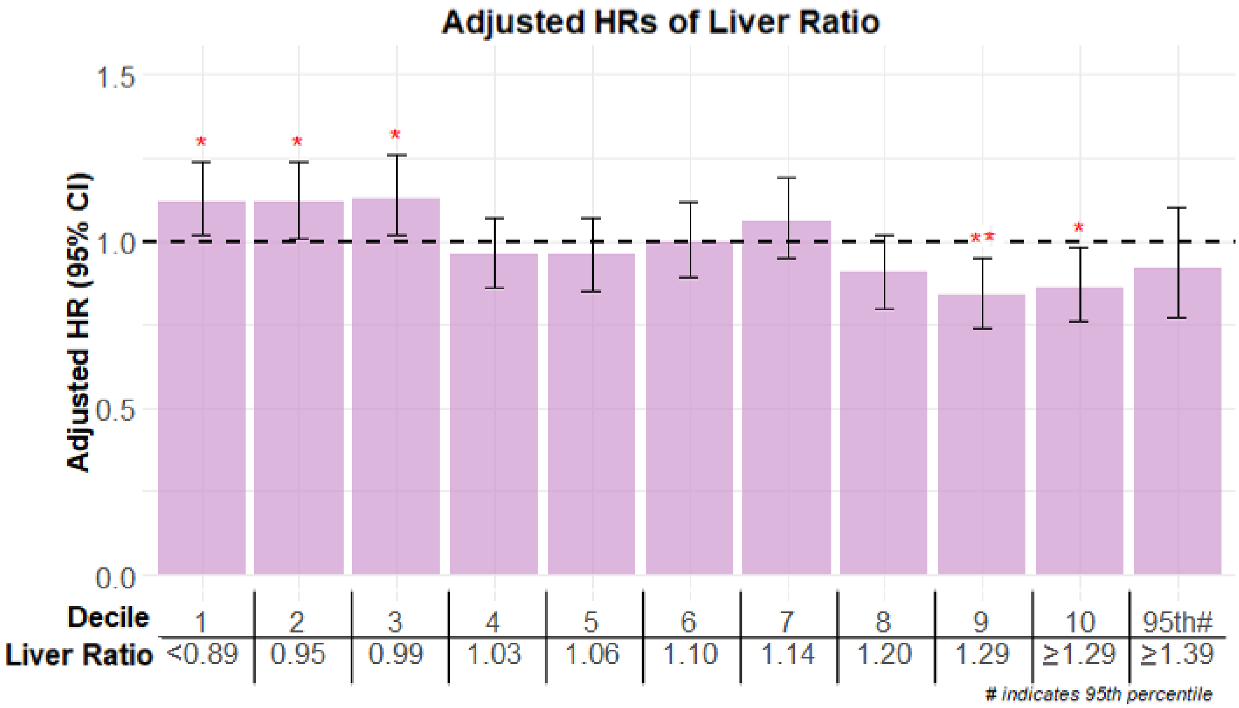


**Supplemental Figure 4. Adjusted hazard ratios for major adverse cardiovascular events (MACE) across deciles of stress/rest liver uptake**. Adjusted hazard ratios for major adverse cardiovascular events (MACE) across deciles of the liver ratio (stress/rest liver uptake) in 10,913 patients undergoing [$^{82}$Rb]Cl PET. A non-linear relationship was observed, with elevated risk in both the lowest (deciles 1–3) and highest (deciles 9–10) liver ratio groups, suggesting bidirectional prognostic associations.

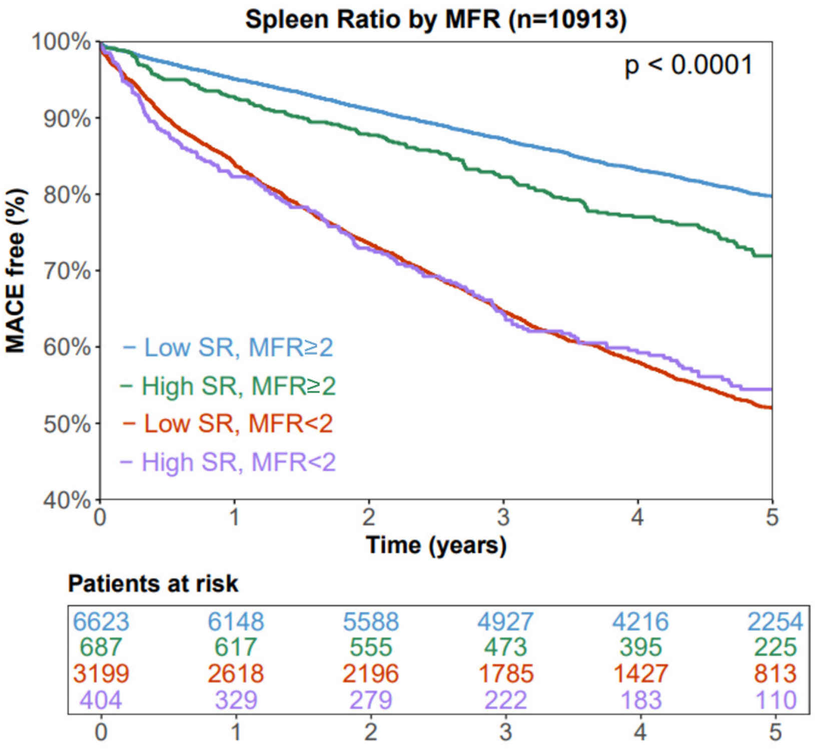


***Hazard Ratios for MACE by SR and MFR Categories***

| SR and MFR Category | Unadjusted HR (95% CI)[1] | Adjusted HR (95% CI)[1] |
|---|---|---|
| High SR, MFR≥2 | 1.41 (1.22 – 1.63)*** | 1.41 (1.22 – 1.64)*** |
| Low SR, MFR<2 | 2.79 (2.59 – 2.99)*** | 1.79 (1.66 – 1.94)*** |
| High SR, MFR<2 | 2.57 (2.21 – 2.98)*** | 1.80 (1.55 – 2.10)*** |

[1]In reference to Low SR, MFR≥2 group

*** p<0.001

**Supplemental Figure 5. Five-Year MACE-Free Survival by Combined Splenic Ratio and Myocardial Flow Reserve Categories.** Kaplan–Meier curves demonstrate cumulative MACE-free survival stratified by splenic ratio (SR) and myocardial flow reserve (MFR). Patients were grouped into four categories: low SR/high MFR (reference group), high SR/high MFR, high SR/low MFR, and low SR/low MFR. Event-free survival was highest in the low SR/high MFR group and progressively decreased across the other strata. The table below the plot provides unadjusted and adjusted hazard ratios (HR) for MACE, using the low SR/high MFR group as the reference. Adjusted HRs were derived from multivariable Cox models. All comparisons were statistically significant ($p < 0.001$).

| Demographic/Clinical Variable | Missingness N (%) |
|---|---|
| Race | 624 (5.72) |
| Smoker | 97 (0.89) |
| Hypertension | 97 (0.89) |
| Dyslipidemia | 97 (0.89) |
| Diabetes Mellitus | 19 (0.17) |
| Family History | 307 (2.81) |
| BMI | 11 (0.1) |
| Previous CAD | 3 (0.03) |
| Past MI | 3 (0.03) |
| Past PCI or stents | 3 (0.03) |
| Past Bypass surgery | 3 (0.03) |
| Rest LVEF | 6 (0.05) |
| Stress LVEF | 7 (0.06) |
| Rest LVEF < 40% | 6 (0.05) |
| Positive stress ECG | 7 (0.06) |

**Supplemental Table 1. Missingness in Demographic/Clinical Data**

The number of missing values we have for the variables mentioned in the paper. All variables with missing data have less than 6% missingness. All variables mentioned in this paper not listed in the table signify that all values were available.

Abbreviations: BMI – body mass index; CAD – coronary artery disease; LVEF – left ventricular ejection fraction; MI – myocardial infarction; PCI – percutaneous coronary intervention; PVD – peripheral vascular disease
